# *hART*: Deep Learning-Informed Lifespan Heart Failure Risk Trajectories

**DOI:** 10.1101/2023.09.04.23295033

**Authors:** Harry Moroz, Yue Li, Ariane Marelli

## Abstract

Heart failure (HF) results in persistent risk and long-term comorbidities. This is particularly true for patients with lifelong HF sequelae of cardiovascular disease such as patients with congenital heart disease (CHD). We developed *hART* (heart failure Attentive Risk Trajectory), a deep-learning model to predict HF trajectories in CHD patients. *hART* is designed to capture the contextual relationships between medical events within a patient’s history. It is trained to predict future HF risk by using the masked self-attention mechanism that forces it to focus only on the most relevant segments of the past medical events. To demonstrate the utility of *hART*, we used a large cohort containing healthcare administrative data from the Quebec CHD database (137,493 patients, 35-year follow-up). *hART* achieves an area under the precision-recall of 28% for HF risk prediction, which is 33% improvement over existing methods. Patients with severe CHD lesion showed a consistently elevated predicted HF risks throughout their lifespan, and patients with genetic syndromes exhibited elevated HF risks until the age of 50. The impact of the birth condition decreases on long-term HF risk. The timing of interventions such as arrhythmia surgery had varying impacts on the lifespan HF risk among the individuals. Arrhythmic surgery performed at a younger age had minimal long-term effects on HF risk, while surgeries during adulthood had a significant lasting impact. Together, we show that *hART* can detect meaningful lifelong HF risk in CHD patients by capturing both long and short-range dependencies in their past medical events. Our code is available at https://github.com/li-lab-mcgill/hART-heart-failure-Attentive-Risk-Trajectory.

## Introduction

The delivery of care for chronic illnesses has become increasingly complex as patients age and their medical histories expand (1). The lifetime risk of heart failure (HF) ranges from 20-45% after the age of 45 years in the United States (2). With advances in medical and interventional therapies, patients with congenital heart disease (CHD), the most common birth defect (3) are now surviving to adulthood (4). In patients with CHD, we have shown that HF is the leading cause of death with a cumulative incidence rate of 25% by age of 65 (5, 6). Adult CHD (ACHD) patients face potential lifelong comorbidities. The demographic changes in the CHD population has motivated a paradigm shift in disease management with an increasing focus on a lifespan perspective and a proactive approach to care (7–9). This shift has refocused attention on the study of the comorbidities that affect the disease outcomes of ACHD. Improved management of comorbidities such as HF are pivotal to improving health outcomes and optimizing the associated healthcare resource utilization for ACHD patients (9–11). While we and others have contributed to understanding the prevalence and risk factors of HF in CHD (12–14), there remains the need to better understand and measure the long-term trajectory of disease progression in this patient population to optimize the timing of medical interventions and available resources (15). Incorporating large datasets to optimize the precision delivery of care, disease trajectories can provide a framework to assess the impact and relationship of different patient presentations, time-varying risk factors and comorbidities on the evolving risk of specific outcomes, such as heart failure.

Assessing the evolving risk of HF in patients with CHD remains a challenging task due to the complexity and the heterogeneity of patient presentations and difficulties in capturing long-range dependencies among variables (5, 12, 13). For many chronic illnesses, such as CHD, modelling the high-dimensional irregular time-varying nature of Electronic Health Record (EHR) data has been limited in its capacity to capture temporal patterns of the disease progression (5, 13, 16).

However, recent advancements in deep learning (DL) have provided highly flexible frameworks that can accurately model long-term sequential patient data, including complex longitudinal clinical and epidemiological presentations of chronic diseases such as CHD (17, 18). Although DL techniques have been shown to be effective in modeling the trajectory of HF in lifespan CHD patients (18), capturing long-term temporal patterns of CHD and generating interpretable trajectories continue to pose a challenge. Models that use Transformer architecture such as Bidirectional Embedding Representations by Transformers (BERT) and Generative Pre-trained Transformer (GPT)-3 (19) (i.e., the same framework used in the recently popularized ChatGPT) have produced remarkable results in natural language processing (NLP) tasks and outperformed recurrent neural networks in modeling sequential data (20–22). This is primarily because Transformers can capture long-and short-range temporal dependency by computing attention between all pairs of time points in an entire sequence (21, 23).

In the context of our study, it is crucial to model the evolving chronic comorbidity conditions caused by CHD (which develops at birth) throughout a patient’s lifespan, requiring the inclusion of the patient’s complete medical history for accurate prediction of disease progression (24). Additionally, from an interpretability standpoint, one motivation for using attention-based learning to model patient medical data is that it allows for a more comprehensive and contextual understanding of the data. Just like how the GPT learns the contextual information within a sentence (19), medical events may have different importance and meaning within a patient’s medical history. Attention-based models can use their attention mechanisms to focus on specific aspects of a patient’s medical history and other relevant data, allowing them to consider the context and individual characteristics of the patient. This can provide a more accurate and nuanced understanding of the patient’s health and can be used to improve treatment planning and decision making. By considering the full context of a patient’s medical history, attention-based models can make more informed and personalized predictions or recommendations, which can ultimately lead to better patient outcomes. Several studies have investigated the use of self-attention in deep learning for patient representation learning, clinical decision support, and predicting future clinical events, and have shown promising results (23, 25–27).

In this study, we propose an attention-based deep learning model called heart failure Attentive Risk Trajectory (*hART*), using longitudinal EHR data to accurately predict lifespan HF risk. Using a longitudinal Quebec CHD dataset containing the administrative medical records for over 130,000 CHD patients with 35 years of follow-up, we evaluated *hART*’s performance in predicting future HF risk in comparison with other methods. We also assess *hART*’s ability to differentiate HF trajectories across populations based on CHD severity, genetic syndromes, and age of death. Furthermore, we explore *hART*’s utility in evaluating individual patients’ HF risk by analyzing the impact of specific medical events over time. Finally, we examine the benefits of performing a pre-training on predicting all medical events followed by fine-tuning on the HF risk prediction task.

## Results

### hART model overview

*hART* uses self-attention to model sequential medical histories of patients to accurately predict their HF risks (Fig. 1). Briefly, we segmented the patient medical records into 6-month intervals and treated them as the time tokens. We utilized transformer layers to model the dependencies among medical events through computing the attention scores between all pairs of time points. To predict the future HF risk in the next 6-month interval, we use the masked self-attention mechanism that forces *hART* to observe only the past medical events. The context vector and attention weights produced by the trained model allow for the interpretation of how medical events contribute to HF risk in the next time interval.

**Figure 1:**
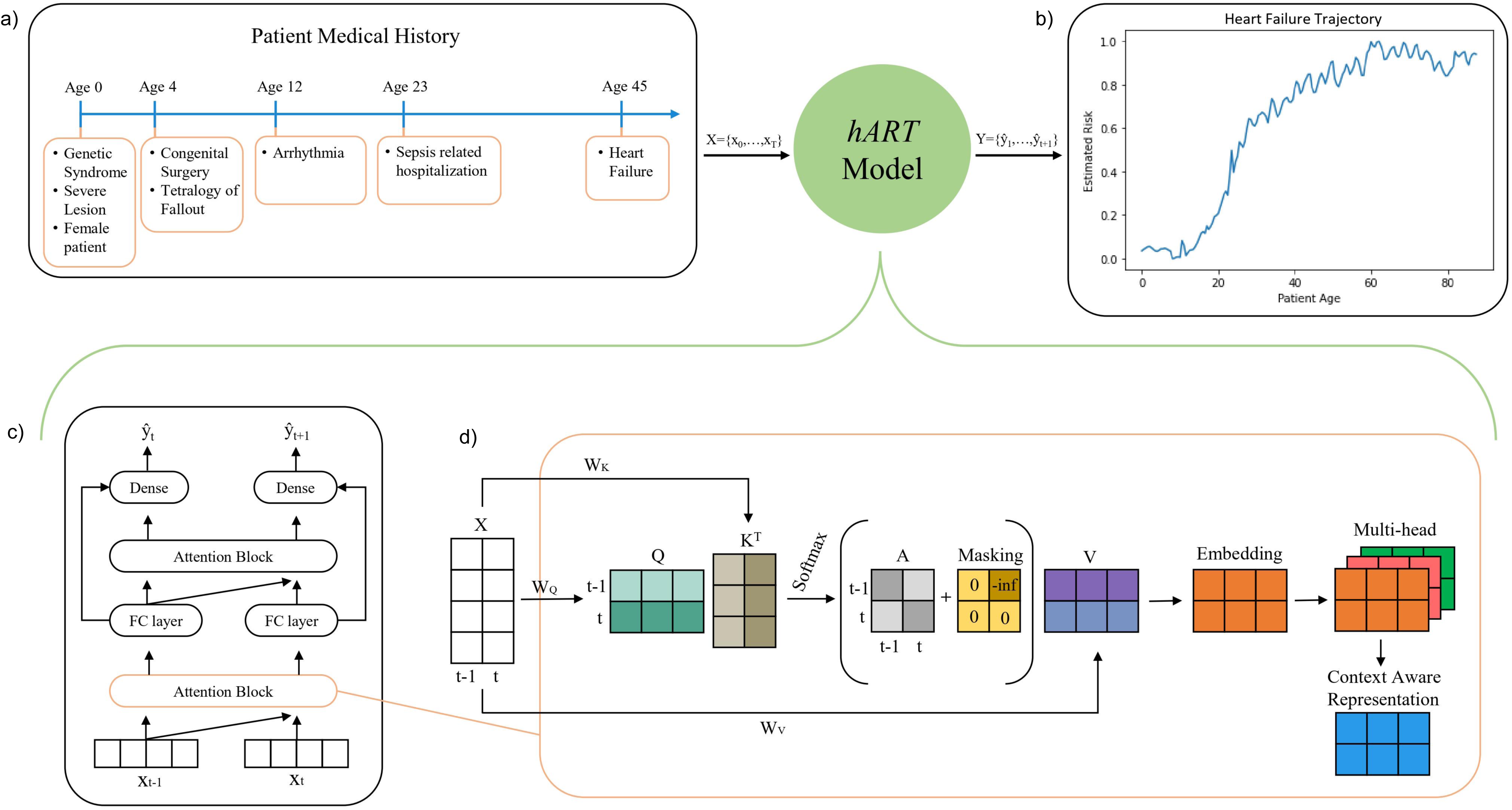
*hART* method overview. (a) A conceptual illustration of the input longitudinal record of a CHD patient. (b) A retrospective patient HF risk trajectory output by *hART*. (c) Attention-based architecture of *hART*. Timesteps at t-1 and t are passed through *hART* to output the predicted values of the future 6-month HF risk, represented as y_t_ and y_t+1_. Each input timestep is passed through two attention blocks connected by a fully connected layer, then a final dense layer that allows the model to output a single value per input. The model also has a skip connection around the second attention block. (d) Masked multi-headed self-attention mechanism of the attention block to prevent the model from peeking into the future.

### HF Prediction Performance

In terms of predicting the next HF event at time t + 1 based on the sequence up to tme t, *hART* achieved an AUPRC score of 0.282 and an AUROC score of 0.967, which outperformed the baseline models (Fig. 2). Particularly noteworthy, it outperformed our previously established GRU-based model, which attained an AUPRC score of 0.210 and an AUROC score of 0.961. The improvements are mostly pronounced at the 1% recall rate, where our model achieved over 80% precision outperforming the other models by 10%. This is promising given the high class imbalance, where HF instances account for only roughly 1% of time steps. The attention model trained on the randomized time steps conferred an AUPRC score of 0.2097 and an AUROC score of 0.9563, which are much lower compared to the *hART* trained on the original time step sequence. This implies that *hART* can leverage the temporal order to improve HF prediction.

**Figure 2.**
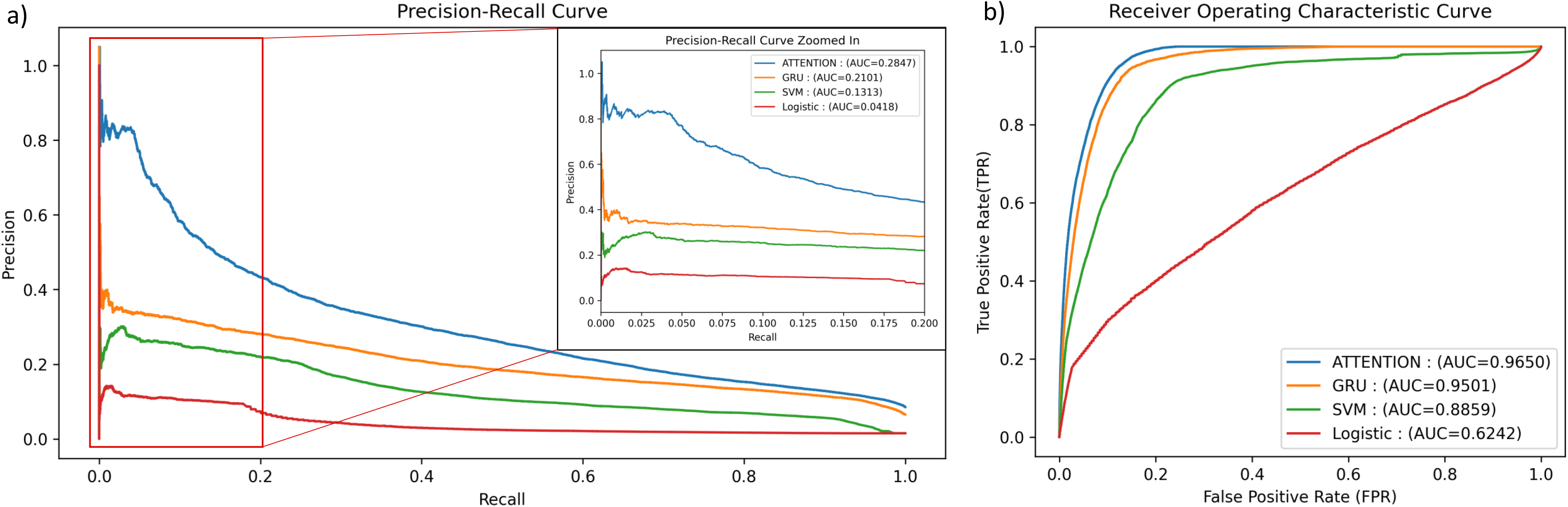
Performance of *hART* and the baseline models in predicting HF. To evaluate each method, we flattened the predicted HF probabilities and the groundtruth binary HF events across all time points and all patients. We then constructed (a) precision-recall curves and (b) ROC curves and computed the AUPRC and AUROC (inside legend) for each method.

### HF Trajectories correlates with CHD Severity

From the data alone, we observe significantly higher estimated risk of HF for patients with severe CHD compared to those with non-severe CHD throughout their entire lifespan (p <0.05, KS-test). Specifically, out of 124,898 patients with non-severe CHD, 11.56% had HF, whereas out of 9,548 patients with severe CHD, 14.04% had HF (p <0.05, Fisher’s exact test). *hART* effectively predicted the actual distribution of HF among severe patients, with the predicted HF trajectory indicating a significant rise in risk 10 years earlier than the actual incidence of HF (Fig. 3a top row) in contrast to the observed frequencies (Fig. 3a bottom row). The highest risk in severe CHD patients begins at 30 years of age, while the incidence of HF rises at 40 years of age. The computed attention matrix for patients with severe CHD highlights the most important time steps for risk prediction, with high attention given to the time of birth and the period from 10 to 40 years of age (Fig. 3a middle row).

**Figure 3:**
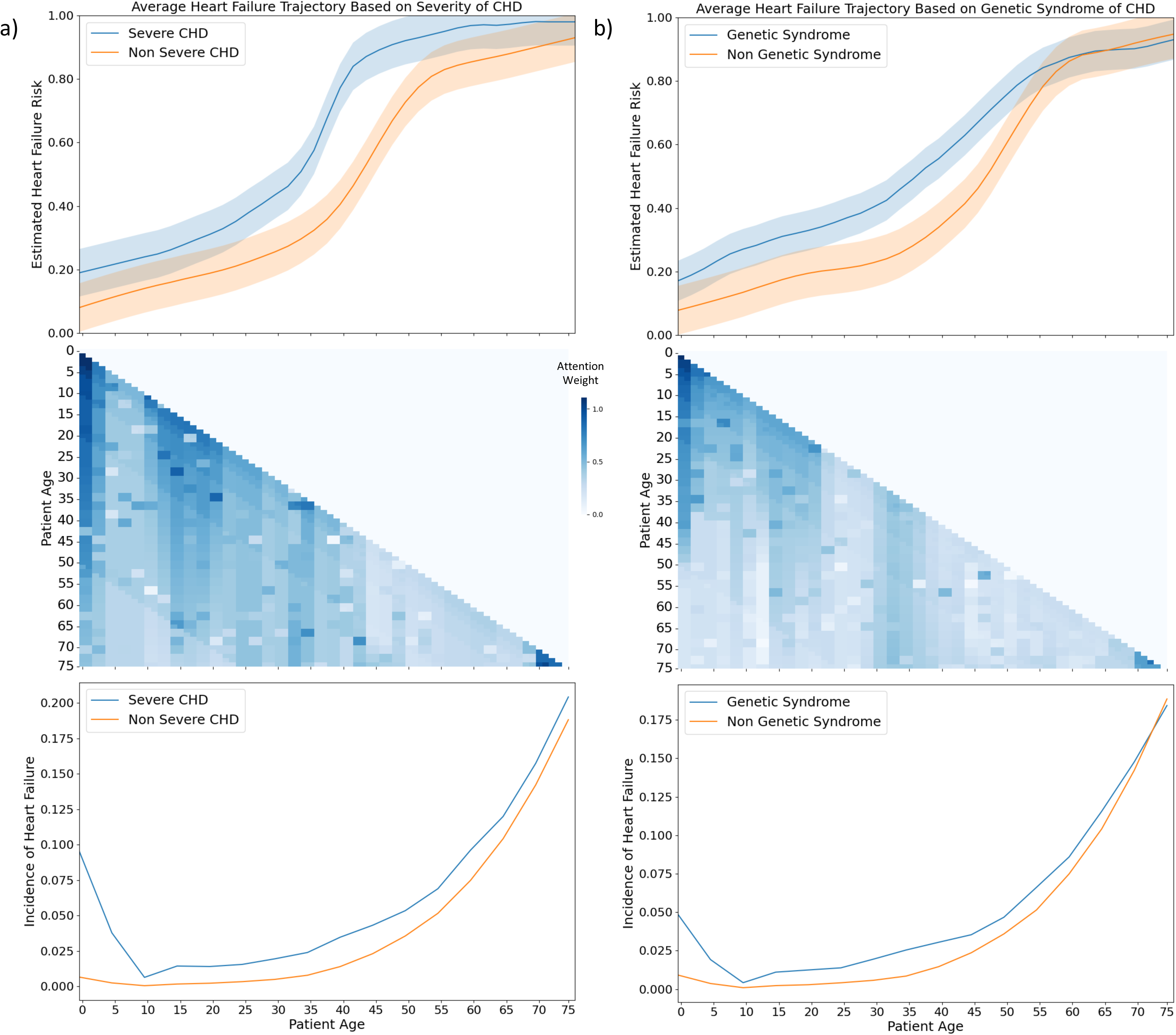
Population-level HF trajectories. The top row depicts a comparison of the predicted HF trajectory between the population with and without a specific attribute (i.e., (a) severe CHD lesion and (b) genetic syndrome). The trajectories are shown as the mean trajectory and the shaded area is the confidence interval of all patients from each population. The middle row depicts the average of attention weight matrix computed over the exposure population, with darker blue representing the time-step, which contributes more to the HF risk prediction at the given time-step (y-axis row). The bottom row depicts the observed frequency of the HF occurrence among the CHD patients.

### CHD patients with high predicted HF risk tend to have genetic syndrome

We first analyzed the impact of genetic variants associated with CHD on the risk of HF using our *hART* model. Patients with known genetic syndrome associated with CHD have a higher risk of developing HF until the age of 50 (p < 0.05, KS-test), after which the risk trajectories intersect with those of non-genetic patients (p > 0.05, KS-test). The comparison between predicted HF trajectories by *hART* and actual HF distribution in genetic and non-genetic patients reveals similar deviations in both groups, with *hART* predicting the intersection of HF occurrence earlier (Fig. 3b top row) compared to the observed frequencies (Fig. 3b bottom row). This suggests that the predictive model is effective in identifying the HF risk in genetic patients more than 10 years earlier than the actual occurrence. Based on the attention heatmap (Fig. 3b middle row), the most important timesteps for predicting HF in genetic patients were found to be from birth to 20 years of age, with decreasing importance until age 50. Interestingly, the initial condition became less important for rises in risk over time, which is supported by the intersection of risk mentioned above. Our *hART* model demonstrates the ability to identify early rises in HF risk in genetic CHD patients and highlights the importance of early interventions in this patient population.

### Predicted HF trajectories correlate with early-age mortalities

We also examined the correlation between increased risk of mortality and HF using the raw data. Our CHD dataset contains 2598 patients who died before 40 years old, 4031 patients who died between 40 and 70, and 11541 patients who died after 70, of which 19.25%, 43.81% and 56.44%, respectively, had HF (p<0.05, Fisher’s exact test). By taking the reciprocal of the predicted HF risk and scaling the trajectory with respect to the HF risk at age of death, we can illustrate the depreciation of overall health (Fig. 4a). First, we find that the initial medical condition of patients who die younger is associate in a lower initial predicted overall health. Secondly, *hART* model predicts that the estimated risk of HF for CHD patients who die younger have significantly earlier decreases in overall heath in predicted HF risk. As expected, the average age of the first HF was lower for patients who had earlier death (p-value < 0.05, KS test). Furthermore, the patient who died before 40 had a significantly higher proportion of surgeries (19.71% vs 9.90% for patients who died between 40 and 70 vs 5.09% for patients who died after 70) (p < 0.05, Fisher’s exact test). By looking at the attention matrix of our population who died before 40, we find that the patient history from birth to the age of 10 contribute significantly to the rapid depreciation of overall health. Although the indication of death was not included in the training of *hART*, we identified a strong association between predicted HF risk and mortality, indicating the potential use of *hART* as a tool for mortality risk stratification and survival analysis in CHD patients.

**Figure 4:**
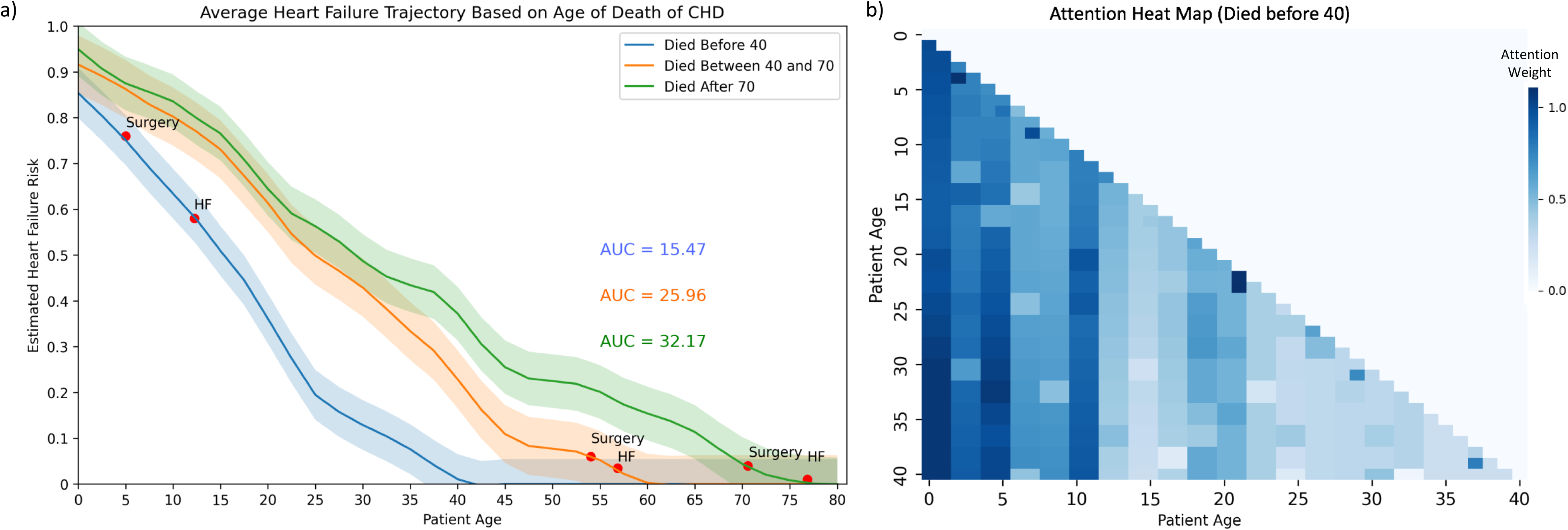
Trajectories based on age of death. (a) Estimated HF risk as a function of patient age. The overall health was calculated by scaling the reciprocal of predicted HF risk by the reciprocal of the observed HF frequency at each age of death. The trajectories reflect the HF-related depreciation of the overall health. (b) The attention weights associated to the population of patients, who died before the age of 40.

### Connecting the dots backward: using the event-based attention to associate HF hospitalization with past medical events in the CHD patient records

We investigated the impact of the timing of medical events on the individualized HF risk trajectories for two representative CHD patients (Fig. 5). Patient A had arrhythmic surgery at the age of 33, and their risk trajectory deviated from the baseline after the surgery. The attention scores computed by *hART* indicated that the surgery remained important to the HF trajectory for the rest of the patient’s life, contributing to the deviation from the baseline (Fig. 5a). Patient B had arrhythmic surgery at the age of 3, and their risk trajectory had a significant deviation from the baseline from the age of 4 to 45. However, *hART*’s attention scores showed that the importance of the surgery in predicting HF decreased over time, and its impact was minimal around the age of 45 (Fig. 5b). These findings suggest that *hART* can capture the understanding of how the context and timing of past medical events affect the lifespan trajectories of individual patients. Such knowledge can be beneficial in designing personalized treatment regimens for CHD patients.

**Figure 5:**
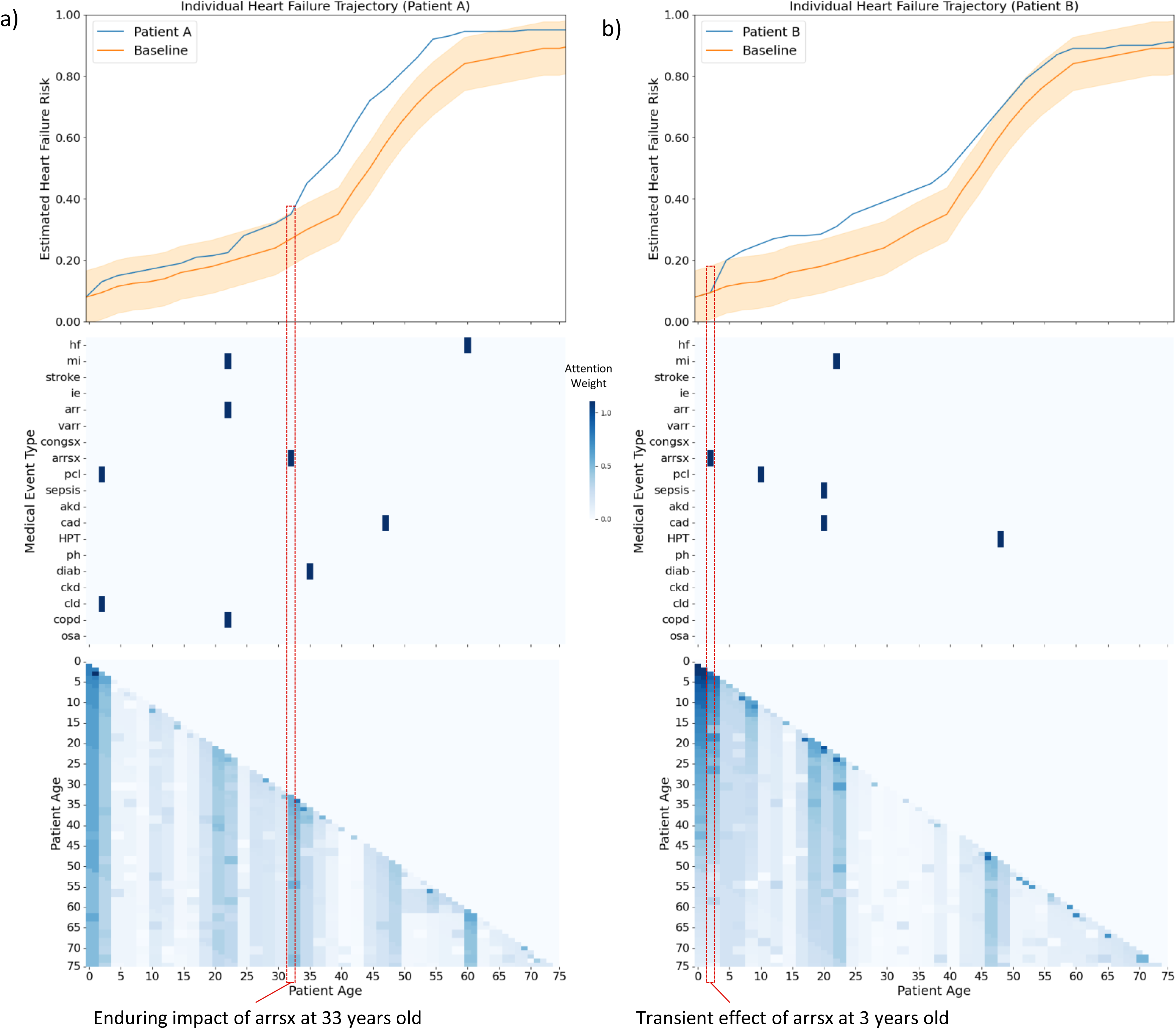
Example of individualized trajectories. For the two example patients in panel a (left column) and panel b (right column), the top row depicts a comparison of the predicted HF trajectory for the selected individual patient and the baseline. The middle row depicts the individual patient’s medical history, with a dark line representing the occurrence of a specified medical event at the given time. The bottom row illustrates the attention weight matrix computed by *hART* for the individual patient. The red boxed illustrate the timing of arrthmic surgery, highlighting the enduring impact of this event at age 33 (Patient A) compared to the transient effect when it occurred at age 3 (Patient B) The baseline trajectory is the mean trajectory and the confidence interval of all patients from the sub-population which Patient A and B belong (i.e. Non-severe CHD lesion, Non-genetic Female Patients with CHD).

### Pre-training on future comorbidities and fine-tuning on HF prediction

We then evaluated the performance of our extended attention-based model, *hART-GPT*, in predicting medical events for CHD patients. We compared the attention-bsaed pre-training with the GRU-based pretraining performances. The attention-based outperformed the GRU-based model for predicting each clinical variable, and was particularly effective at predicting comorbidities associated with HF, such as stroke, infective endocarditis, sepsis, myocardial infarction, and acute kidney disease. Additionally, we examined the performance of fine-tuning *hART-GPT* on predicting the next HF and compared it to *hART* and other baseline models (Fig. 6b). We observed that *hART-GPT* slightly outperformed *hART* with an AUROC of 0.979 and an AUPRC of 0.2900. These results suggest that *hART-GPT* is a promising tool for predicting the risk of HF and associated comorbidities in patients.

**Figure 6:**
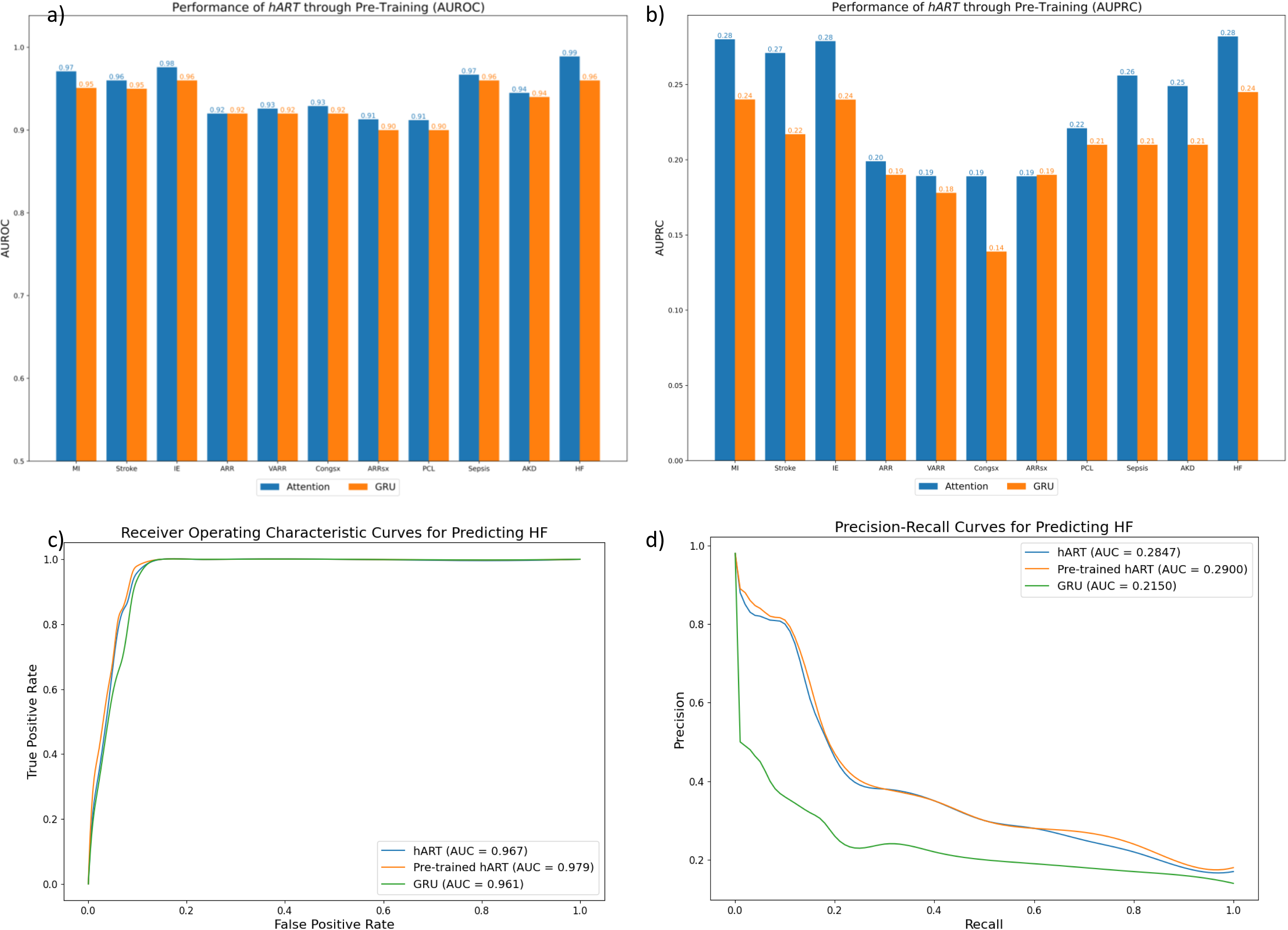
*hART*-*GPT* performance. (a) and (b) AUROC and AUPRC for predicting each of the 11 time-varying variables between the attention-based *hART-GPT* model and the GRU-based model. (c) and (d) ROC and PRC curves comparing between *hART-GPT* and *hART*.

## Data Availability

All data produced in the present work are contained in the manuscript.

https://github.com/li-lab-mcgill/hART-heart-failure-Attentive-Risk-Trajectory

## Acknowledgment

Harry Moroz is funded by FRQ-S Master’s Training Award. Dr. Yue Li is supported by a NSERC Discovery Grant and Canada Research Chair (Tier 2) for Machine Learning in Genomics and Healthcare. Dr. Ariane Marelli is supported by the Heart and Stroke Foundation grant and award (John Day, MD, Excellence Award for ’Heart Failure Trajectory Along the Care Continuum for Congenital Heart Disease Patients Across the Lifespan).

## Discussion

HF trajectories enable the assessment of the progression of chronic diseases and comorbidities throughout a patient’s life. By analyzing long-term sequential patient data, this approach can provide insight into the evolution of a disease over time, including the impact of several factors such as age, sex, and individual patient characteristics. This valuable capability holds the potential to inform precision delivery of care, leading to improved patient outcomes and optimized resource allocation for individuals with CHD (9). In this study, we present the first use of attention-based deep learning in predicting HF among the Quebec CHD patient cohort. Our developed approach called heart-failure Attentive Risk Trajectories (*hART*) is a step towards a deep understanding of the lifespan disease progression of CHD, and specifically the leading comorbidity, HF. We demonstrate the capacity of *hART* on modeling sequential administrative diagnostic data from Quebec CHD patients and its ability to accurately predict their HF trajectories. The remarkable performance of *hART* compared to existing approaches demonstrates the efficacy of the attention mechanism in effectively modeling long-term sequential patient data. It allows for contextualization and focused attention on specific medical events within a patient’s comprehensive medical history. Moreover, we showed that the temporal order is important to achieve the highest prediction accuracy of *hART*, implying its ability to learn hidden semantics from the sequential medical data. Additionally, we showed the potential of training a foundational Generative Pre-training Transformer (GPT) framework (*hART-GPT*) by first predicting the associated comorbidities in patients and then fine-tuning to further improve predicting the HF risk. Our model can be used to tackle a wide array of medical problems beyond HF (23).

Furthermore, we demonstrate the use of predicted HF trajectories to gain a deeper understanding of patient’s HF risk and medical profile. Our model has captured how different medical events and patient population of CHD patients can lead to different level of long-term HF risk.

Assessing HF trajectories based on population reveal meaningful clinical implications. Firstly, despite severity not being included in the training of our model, *hART* identified that patients with severe CHD exhibited consistently elevated risks of HF throughout their lifespan, which supports recent studies(14). Our results suggest that future studies can use the model to effectively stratify patients into risk groups, which could inform the allocation of resources towards patients with higher severity. Secondly, our findings provide evidence that the predicted HF trajectory for patients with genetic CHD aligns with previous studies indicating a higher likelihood of surgery, complication and HF in this specific cohort (37). We observe that once patients with genetic syndrome reach the age of 50, their risk profile starts resembling that of patients without the genetic syndrome. Moreover, by utilizing the attention matrix heatmaps, we can identify the diminishing influence of early time steps in patients with genetic syndrome over time. These results highlight the ability of *hART* to understand the significance of birth conditions and their evolving impact on HF throughout individuals’ lifespans, which is crucial in managing congenital and chronic diseases. Thirdly, our results support the correlation between the evolving risk of HF and mortality, indicating that the model can accurately capture this relationship despite age of death not being included in the training (38). This motivates the use of HF trajectories to assess the overall lifespan health condition of patients with CHD and to inform clinical decision-making and the timing of specialty referral (6, 12). Finally, our study showcased how *hART* can assess HF risk for individual patients and can capture the importance of the timing of medical events, such as arrhythmic surgeries. Our findings underscore the attention mechanism’s ability to contextualize medical events within patient histories.

This study also provides one of the first visualizations of lifelong HF trajectories, providing a novel tool for clinicians to use the power of deep learning to inform their interventions (9, 15). Comparing patient populations can help clinicians understand the trends and impacts of patient histories on lifespan HF risk. Additionally, the use of HF trajectories at an individualized level can facilitate the precision delivery of care by identifying turning points and transitions periods based on the process of health development and health determinants such as medical events and genetic predisposition (9, 39, 40). The attention mechanism in *hART* provides an advantage over previously used RNN models (18) by offering interpretability for clinicians, allowing for a clear understanding of the factors, specifically how the timing of medical events contribute to the model’s predictions (26). The addition of the informative attention weights can provide valuable information to clinicians about the most important medical events in a patient’s life. By visualizing the attention weights at each time point, we can show which data points the model is focusing on and the relative importance of each data point. This can help clinicians identify key medical events, such as the onset of a disease or the start of a treatment and understand their impact on the patient’s overall health.

Future research in the field of CHD will focus on analyzing the capabilities of deep learning-informed disease trajectories to better understand the complex nature of lifespan diseases, incorporating a wide range of medical data and assessing different outcomes (9). While the current model shows promise with its flexibility and generalizability, there are limitations to be addressed. One limitation of modeling patient data in discretized time intervals using attention is that it may not capture the full complexity of the data. In many cases, medical data is recorded at irregular intervals, which can provide deeper insight into the progression of a disease (41).

Additionally, discretizing the time intervals may make it more difficult to model the autoregressive nature of the data, where future time points are predicted based on previous ones. Another limitation which we will address in future studies is the inclusion of only 23 selected variables. The flexibility and generalizability of *hART* will facilitate the inclusion of more patient features, and assessment of different outcomes in future studies. In terms of clinical applications, the success of our model will motivate its use to analyze other clinical questions such as, finding the optimal time for intervention for an individual patient, stratifying patients into risk groups based on trajectories, and examining the impact of specialist follow-ups on the quality of life of CHD patients (15). For example, one way to find the optimal time for follow-up using predicted disease trajectories is to identify the points in the trajectory, where there is a significant change in the risk of the disease or in the patient’s overall health. These points may indicate a need for additional medical attention or intervention and can provide guidance on the timing of follow-up visits. Moreover, comparing the predicted trajectories of different patients can provide insight into common patterns and trends, which can inform the development of general follow-up guidelines. By using predicted disease trajectories in combination with clinical expertise, it is possible to find the optimal time for follow-up and ensure that the right patient receives the right care at the right time (6, 9).

In conclusion, the deep learning framework of lifelong HF trajectories provided by this study empower clinicians to effectively identify high-risk individuals, optimize intervention timing, and assess the evolving impact of comorbidities in CHD patients. Overall, this study highlights the value of disease trajectory modeling in improving our understanding of the long-term effects of CHD complications.

## Methods

### Data source

In Quebec, Canada, a unique Medicare number is assigned to individuals in their first year of life, enabling comprehensive tracking of diagnoses and health services throughout their lifetime. This study utilized administrative databases, including physicians’ services and drug claims, hospital discharge summaries, and the Quebec Health Insurance Board and Death Registry.

These databases contain vital statistics, demographic information, and diagnostic and procedure codes (ICD-9 and ICD-10) recorded since 1983. The Quebec CHD Database, one of the largest of its kind worldwide, was created by merging and cleaning data from these sources, identifying CHD patients using validated algorithms and optimizing the accuracy of diagnoses through manual audits (4).

The resulting dataset is from the Quebec CHD database, comprising 137,493 patients with up to 35 years of follow-up from 1983 to 2017. It includes demographic information, inpatient and outpatient diagnoses, surgical history, and vital status. HF events were identified based on hospitalizations with heart failure (HFH) as the admission and/or discharge diagnosis. The dataset includes demographic variables (sex and age), diagnoses (CHD lesion type, pulmonary hypertension, and coronary artery disease), hospitalizations (acute kidney injury and sepsis), and surgeries (percutaneous procedures, congenital surgeries, and surgical arrhythmia procedures). These variables were selected based on their relevance as potential predictors and common confounding factors associated with HF.

### Study population

The study population cohorts were obtained from the Quebec CHD database. For the training and evaluation of our models, we applied two exclusion criteria. Firstly, patients who entered the study after the age of 75 were excluded. Secondly, patients with less than 6 months of available data were excluded. Following these criteria, a total of 134,446 patients were included in the training and evaluation of our models. Out of these patients, 11.74 % experienced at least one HFH with the average age of HF being 62.97.

### EHR data processing

We utilized a sequence-to-sequence supervised approach to predict future HF risk and generate a continuous lifespan trajectory of HF risk. Patient records were partitioned into six-month intervals, with each interval comprising a 23-dimensional vector encompassing static variables reflecting the patient’s condition at birth (such as sex and genetic syndrome), a continuous variable (age), and binary-encoded indicators denoting the occurrence of particular medical events and medical conditions (such as surgeries and comorbidities). We opted for 6-month intervals to enhance the representation of medical events and positive HF labels during model training, while also maintaining consistency with our earlier developed models (18). The labels were generated using a 1-step sliding window technique based on HF hospitalization. This approach enabled us to predict the future 6-month risk of HF at each time step, creating a continuous prediction of HF risk based on past patient data. To ensure consistency, patient sequences were masked with values of -1, resulting in all sequences being standardized to 150 time-steps, which corresponds to the maximum age of 75 years.

### hART (heart failure Attentive Risk Trajectory Model)

Our *hART* has four major components — *input padding*, *positional encoder, masked multi-headed self-attention layers, and residual connections*:

1. Firstly, padding is applied to each patient to extend all inputs to the size corresponding to the oldest age of any patient in the dataset, and masking is used to ignore these special tokens in loss calculation and predictions.
2. Secondly, in contrast to approaches like RNNs, the attention mechanism does not utilize any recurrence or convolutions for sequence modeling. Instead, we pass the data through a positional encoder to translate the notion of time within the time steps of our sequences (21). The positional encoder is commonly used in NLP problems where the transformer is employed. We chosen to use the same discrete positional encoder as described in the original Transformer paper (21), however, Time2Vec (30), a trainable encoding layer, was also tested and yielded inferior results. Positional encoding describes the location or position of an entity in a sequence so that each position is assigned a unique representation.
3. Thirdly, we pass our data through the attention layers. A masked matrix is added to the scaled dot product equation to ensure that the attention weights only consider time points before the predicted time step to be included in the calculation of the context vector (Fig. 1). Each masked multi-head attention mechanism is followed by a normalization layer and a dropout layer to form an attention block. Our proposed model uses two sequential layers of this attention block. This allows the model to build up a hierarchical representation of the input data, with each layer focusing on a different level of abstraction. This can help the model to better capture the complex relationships and dependencies in the data, and can also improve its performance and interpretability (21, 26).
4. Finally, the implementation of a residual connection between the two attention blocks in the model mitigates the vanishing gradient problem during backpropagation. It also provides the output representation with knowledge of the original state of the input. This reinforces the contextual representations of a patient’s medical risk assessment (33).

The output sequence from the attention layers is a context-aware representation of a patient’s health progression. Furthermore, the representation is computed with respect to the future occurrence of HF and thus is associated to the evolving risk of HF of a patient. The final representation is passed through a final dense layer with a sigmoid activation function, computes a single-value HF risk probability estimate for each time-step. This works as a binary classification layer for each time step. The output is a sequence of risk score predictions for each 6-month interval as a deep learning-informed lifespan HF risk trajectory.

As the dataset labels were heavily skewed towards negative examples due to the limited number of recorded HF hospitalizations per patient, we employed focal loss to improve the precision of our model on the imbalanced dataset. Focal loss down-weights well-classified examples and focuses on misclassified examples (18, 36).

### hART-GPT (hART - Generative Pre-Trained Transformer)

*hART-GPT* is an extension of the *hART* architecture described earlier, where we replace the final dense layer to outputs the 11 time-varying variables that make up a patient’s 6-month medical visit. By doing so, we can create a generalized patient representation that can be fine-tuned to predict any outcome, such as heart failure. To this end, we fine-tune the fixed output of the attention blocks of the pre-trained *hART-GPT* by appending a dense layer with one output node to predict the HF variable. This approach allows us to leverage GPT’s ability to predict the next word in a sentence to *hART*’s ability to predict the next medical event. In our case, we are fine-tuning the model to predict the next HF event. This combination allows for a more accurate and personalized prediction of HF risk based on a patient’s medical history, ultimately leading to better patient outcomes.

### Method evaluation

We compared the performance of the proposed model, *hART*, with three baseline models: logistic regression (LR), Support Vector Machine (SVM) and our previously published Deep Heart-Failure Trajectory Model (DHTM) (18), which utilizes Gated-Recurrent Units (GRU). Additionally, to assess the significance of the order of medical events within a patient’s medical history, we have randomized the time steps and re-evaluated *hART* (34). We divided our patient dataset into a training set and a testing set, with 80% used for training and 20% for evaluation. Out of the 80% training set, we further split 15% as a validation set for hyperparameter tuning using the Adam optimizer (35). The optimal parameters for each model are listed in **Supplementary Table 1**. To evaluate the performance of our models, we calculated two metrics: Area Under the Precision Recall Curve (AUPRC) and the Area Under the Receiver Operating Characteristic (AUROC) curve. In our binary classification task, we utilize the optimized threshold described in (18), which is applied to determine whether a predicted outcome should be classified as positive or negative.

### Statistical analysis

We examined the average HF trajectory and confidence interval of patients in different subgroups of CHD and computed the significant difference between the sequences of predicted risk using the Kolmogorov-Smirnov (KS) test. Specifically, we considered 3 types of subgroups: genetic syndrome, severe CHD (encompassing conditions such as tetralogy of Fallot, truncus arteriosus, transposition complexes, endocardial cushion defects, or univentricular heart), and age of death. For unbiased analysis, we computed the HF trajectories on the held-out patients (testing set), which were not used to train *hART*. Alongside the predicted HF trajectories, we visualize the *hART*-generated attention weights for the exposure group as a heat maps. Finally, we compute the distribution of actual occurrences of HF for each subpopulation. Table 1 summarized the characteristics of each population. The Fisher’s exact test is used to determine the significance of differences between characteristics of each patient profile.

**Table 1:** Comparison of selected demographic and clinical characteristics between different CHD patient profiles. The percentage of patients within specific patient profiles who have experienced heart failure (HF) and undergone surgery, as well as the prevalence of certain medical and demographic characteristics. Additionally, the table provides the average age at which medical events occurred within each patient profile. The table is intended to provide insights into the distribution of medical events and patient characteristics within the studied patient profiles, and to facilitate further analysis of patient subgroups.

|  | H<br>F<br>(<br>%)<br>) | HF Age | Surgery<br>(%) | Surgery<br>Age | Female<br>(%) | Severity<br>(%) | Genetic<br>Syn (%) |
| --- | --- | --- | --- | --- | --- | --- | --- |
| **All Patients**<br>(n = 137493) | 1<br>1<br>.<br>7<br>4 | 62.97 | 7.30 | 23.09 | 50.17 | 7.10 | 8.32 |
| **Male**<br>(n = 68514) | 1<br>2<br>.<br>4<br>1<br>* | 61.26* | 7.86* | 23.82 |  | 6.86 | 8.68* |
| **Female**<br>(n = 68979) | 1<br>1<br>.<br>0<br>7<br>* | 64.88* | 6.74* | 22.24 |  | 7.35 | 7.95* |
| **SevereCHD**<br>(n = 9764) | 1<br>4<br>.<br>0<br>4<br>* | 23.22* | 39.54* | 8.16* | 51.89 |  | 21.01* |
| **Non SevereCHD**<br>(n = 127729) | 1<br>1<br>.<br>5<br>6<br>* | 66.66* | 4.84* | 32.41* | 50.04 |  | 7.35* |
| **Genetic Syndrome**<br>(n = 11433) | 1<br>0<br>.<br>8<br>5 | 35.02* | 15.68* | 7.24* | 47.96* | 17.94* |  |
| **No Genetic Syndrome**<br>(n = 126060) | 1<br>1<br>.<br>8<br>2 | 65.30* | 6.54* | 26.53* | 50.37* | 6.12* |  |
| **Age of Death (<40 years old)**<br>(n = 2598) | 1<br>9<br>.<br>2<br>5<br>* | 12.27* | 19.71* | 5.01* | 45.30 | 33.56* | 28.87* |
| **Age of Death (>40 and <70 years old)**<br>(n = 4031) | 4<br>3<br>.<br>8<br>1<br>* | 56.85* | 9.90* | 54.04* | 42.12 | 6.70* | 7.24* |
| **Age of Death (>70 years old)**<br>(n = 11541) | 5<br>6<br>.<br>4<br>4<br>* | 76.84* | 5.09* | 70.56* | 51.10 | 1.44* | 3.51* |

\* Statistically significant at P<0.05.

### Evaluating the performance of *hART-GPT*

We evaluate the performance of the *hART-GPT* model in predicting the 11 medical events which were binary encoded in phase one. These variables include myocardial infarction (MI), Stroke, infective endocarditis (IE), arrhythmia (ARR), ventricular arrhythmia (VARR), congenital surgery (Congsx), arrhythmic surgery (ARRsx), Percutaneous Coronary Intervention (PCI), Sepsis, acute kidney disease (AKD), and HF. The labels for these variables were generated using a 1-step sliding window technique, allowing the model to predict the occurrence of these events within the subsequent 6-month period. To assess the performance of *hART-GPT*, the AUROC and AUPRC for each variable was compared against a GRU-based model (18). Additionally, we compared the AUROC and AUPRC of the fine-tuning of *hART-GPT* against *hART* for predicting HF.

