## Supplementary Table 1 for "*hART*: Deep Learning-Informed Lifespan Heart Failure Risk Trajectories"

Optimal number of attention blocks: 2

Number of attention heads in Attention Block 1: 8

Number of attention heads in Attention Block 2: 8

Hidden dimension size of the encoder layers: 64

Learning rate and weight decay of the optimizer: 0.001

Dropout rate to prevent overfitting: 0.2

Batch size used during training: 128

Maximum sequence length allowed for training and inference: 150

Number of epochs for training the model: 232
